# Chronic Low Back Pain Causal Risk Factors Identified by Mendelian Randomization: a Cross-Sectional Cohort Analysis

**DOI:** 10.1101/2024.09.23.24314235

**Authors:** Patricia Zheng, Aaron Scheffler, Susan Ewing, Trisha Hue, Sara Jones, Saam Morshed, Wolf Mehling, Abel Torres-Espin, Anoop Galivanche, Jeffrey Lotz, Thomas Peterson, Conor O’Neill, REACH investigators

**Author notes:** ^**^Joint first authors (Patricia Zheng, Aaron Scheffler). **CORRESPONDING AUTHOR:** Patricia Zheng, MD, Associate Professor, Orthopaedic Surgery, 1500 Owens St., San Francisco CA 94158.

## Abstract

**Background Context:** There are a number of risk factors- from biological, psychological, and social domains- for non-specific chronic low back pain (cLBP). Many cLBP treatments target risk factors on the assumption that the targeted factor is not just associated with cLBP but is also a cause (i.e, a causal risk factor). In most cases this is a strong assumption, primarily due to the possibility of confounding variables. False assumptions about the causal relationships between risk factors and cLBP likely contribute to the generally marginal results from cLBP treatments.

**Purpose:** The objectives of this study were to a) using rigorous confounding control compare associations between modifiable causal risk factors identified by Mendelian randomization (MR) studies with associations in a cLBP population and b) estimate the association of these risk factors with cLBP outcomes.

**Study Design/Setting:** Cross sectional analysis of a longitudinal, online, observational study.

**Patient Sample:** 1,376 participants in BACKHOME, a longitudinal observational e-Cohort of U.S. adults with cLBP that is part of the NIH Back Pain Consortium (BACPAC) Research Program.

**Outcome Measures:** Pain, Enjoyment of Life, and General Activity (PEG) Scale.

**Methods:** Five risk factors were selected based on evidence from MR randomization studies: sleep disturbance, depression, BMI, alcohol use, and smoking status. Confounders were identified using the ESC-DAG approach, a rigorous method for building directed acyclic graphs based on causal criteria. Strong evidence for confounding was found for age, female sex, education, relationship status, financial strain, anxiety, fear avoidance and catastrophizing. These variables were used to determine the adjustment sets for the primary analysis. Potential confounders with weaker evidence were used for a sensitivity analysis.

**Results:** Participants had the following characteristics: age 54.9 ± 14.4 years, 67.4% female, 60% never smokers, 29.9% overweight, 39.5% obese, PROMIS sleep disturbance T-score 54.8 ± 8.0, PROMIS depression T-score 52.6 ± 10.1, Fear-avoidance Beliefs Questionnaire 11.6 ± 5.9, Patient Catastrophizing Scale 4.5 ± 2.6, PEG 4.4 ± 2.2. In the adjusted models alcohol use, sleep disturbance, depression, and obesity were associated with PEG, after adjusting for confounding variables identified via a DAG constructed using a rigorous protocol. The adjusted effect estimates- the expected change in the PEG outcome for every standard deviation increase or decrease in the exposure (or category shift for categorical exposures) were the largest for sleep disturbance and obesity. Each SD increase in the PROMIS sleep disturbance T-score resulted in a mean 0.77 (95% CI: 0.66, 0.88) point increase in baseline PEG score. Compared to participants with normal BMI, adjusted mean PEG score was slightly higher by 0.37 points (95% CI: 0.09, 0.65) for overweight participants, about 0.8 to 0.9 points higher for those in obesity classes I and II, and 1.39 (95% CI: 0.98, 1.80) points higher for the most obese participants. Each SD increase in the PROMIS depression T-score was associated with a mean 0.28 (95% CI: 0.17, 0.40) point increase in baseline PEG score, while each SD decrease in number of alcoholic drinks per week resulted in a mean 0.12 (95%CI: 0.01, 0.23) increase in baseline PEG score in the adjusted model.

**Conclusions:** Several modifiable causal risk factors for cLBP - alcohol use, sleep disturbance, depression, and obesity- are associated with PEG, after adjusting for confounding variables identified via a DAG constructed using a rigorous protocol. Convergence of our findings for sleep disturbance, depression, and obesity with the results from MR studies, which have different designs and biases, strengthens the evidence for causal relationships between these risk factors and cLBP (1). The estimated effect of change in a risk factors on change in PEG were the largest for sleep disturbance and obesity. Future analyses will evaluate these relationships with longitudinal data.

## Introduction

There are a number of risk factors-from biological, psychological, and social domains- for non-specific chronic low back pain (cLBP) (1). Associations between these risk factors and cLBP underlie the widely accepted conceptual model of cLBP, the biopsychosocial model (2). Clinical guidelines for treatment of cLBP recommend a number of different interventions targeted to risk factors associated with the biopsychocial model, most notably therapeutic exercise (3–5), pain neuroscience education (3), manual therapy (3–5), acupuncture (3–5), and cognitive behavioral therapy (CBT) (5). However, the effects from randomized controlled trials (RCT’s) of these treatments are at best modest (6). One reason why cLBP intervention fail may be that the risk factors they target, while they are associated with cLBP, do not cause cLBP. Targeting treatments to causes may lead to new, more effective, therapeutic approaches.

Determining if a risk factor causes cLBP requires studies that minimize confounding bias (i.e., bias due to variables that are a common cause of both a risk factor and an outcome). There are two general approaches for addressing confounding bias: design-based and analysis-based (7). Analysis-based approaches use statistical methods to minimize bias of estimated associations in observational data by adjusting for confounding variables. A common approach for identifying confounders and their corresponding adjustment sets is to construct a directed acyclic graph (DAG), which embeds existing knowledge and theory into a causal graph describing the relationship among risk factors, outcomes, and other important variables (8). Design-based approaches rely on study design, rather than statistical methods, to address confounding bias. The most robust design-based approach is a randomized controlled trial (RCT). While RCT’s are the gold standard for establishing causality (9) for most cLBP risk factors random allocation is either not possible (e.g., obesity) or unethical (e.g. smoking). An alternative design-based approach increasingly used in cLBP research is Mendelian randomization (MR). MR uses germline genetic variants as proxies for risk factors (9). As genetic variants are randomly assigned at conception, they should be independent of confounding factors (9). Therefore, MR attempts to produce comparisons analogous to an RCT, with individuals randomized to a particular genotype, rather than an intervention (9). When specific assumptions are met, the strength of evidence for MR studies lies somewhere between observational studies and randomized controlled trials (RCT’s) (10). While MR and other design-based methods are important tools they have limitations (7). As design-based and analysis-based methods have different underlying assumptions and biases, triangulating the findings from studies done with both approaches provides stronger evidence for causal links than either method independently (7).

Recent MR studies have identified causal links between a variety of risk factors and cLBP (10–36), many of which are modifiable and therefore potential treatment targets. MR cLBP causal effect estimates are based on the association between genetic variants and prevalent cLBP cases in a population, using large publicly available databases (10). The objectives of this study were to a) compare associations between modifiable causal risk factors identified by design-based MR studies with associations defined by an analytic approach in a cLBP population and b) estimate the association of these risk factors with cLBP outcomes. To accomplish these objectives we used data from a unique cLBP cohort study (BACKHOME) (37), which contains measurements of numerous, heterogenous variables from a large number of participants. Variables in this dataset that have been identified in MR studies as modifiable causes of cLBP were selected as exposures (alcohol use, smoking, sleep disturbance, depression, and obesity (10, 23–25, 38) and associations with a composite outcome of pain intensity and interference (PEG score) were determined. Confounding bias was controlled using statistical adjustment based on factors identified via a DAG constructed using a rigorous and structured protocol, and the magnitude and direction of association synthesized with MR results to identify the potential impact of interventions targeted to these risk factors.

## Methods

### Study Design

Cross sectional analysis of a longitudinal, online, observational study.

### Setting

The study was built on the NIH-supported Eureka Research Platform, which allows for the development and hosting of digital clinical studies. It allows completely remote web- and mobile-based recruitment, enrollment, consent, and participation across the United States (39). Enrollment started in July 2021 and will continue until approximately 3,000 participants have been enrolled. Participants will be followed 2 years or more, with surveys completed every 3 months the first year then every 6 months thereafter. This analysis included data from the baseline survey only, using data collected from 1,868 participants who had enrolled in BACKHOME and completed baseline surveys through April 18, 2023. (Figure 1).

**Figure 1:**
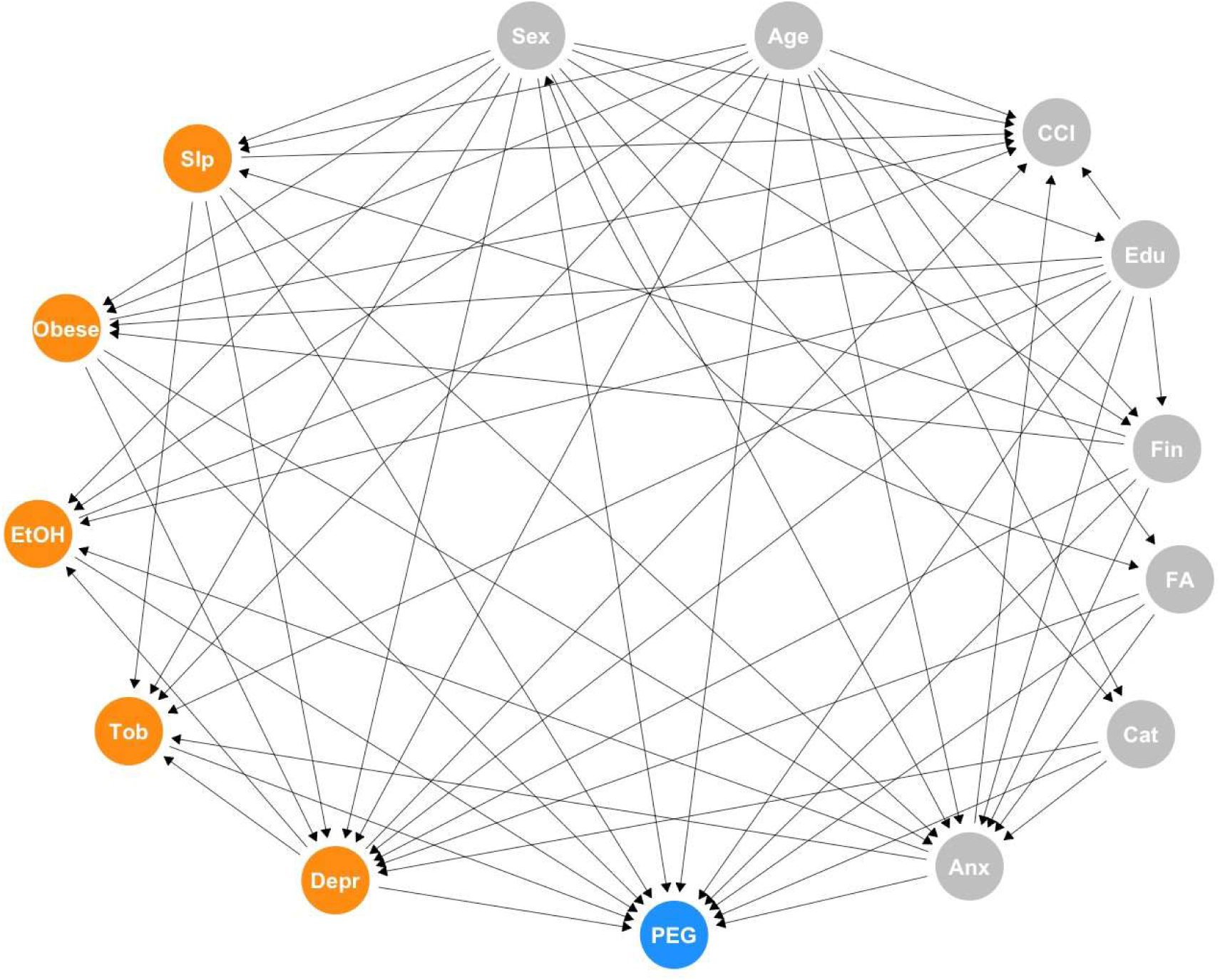
PEG = three-item scale for assessing pain intensity and interference; FA = Fear Avoidance; Cat = Pain Catastrophization; Depr = Depression; Anx = Anxiety; Slp = Sleep; EtOH = Alcohol use; Tob = Smoking; CCI = Charlson Comorbidity Index; Edu = Education; Fin = Financial strain. Exposures highlighted in orange.

### Participants

Participants had to be 18 years of age or older, registered for a Eureka account, currently living in the United States, have an iOS or Android smartphone, have a cell phone number, agree to participate in English, and be able to provide consent to participate in the study. After providing electronic consent to participate in the study, participants were asked to complete a baseline survey about demographics, medical conditions, medications, and behaviors through the study app. Participants could voluntarily provide permission to collect additional data from their smartphones, including geolocation and, among iOS users, HealthKit.

In addition to meeting the requirements for the Eureka platform registration participants had to meet the criteria for cLBP as defined by NIH Pain Consortium Research Task Force (RTF) and BACPAC Minimum Dataset Working Group: current self-report of chronic low back pain (pain between the lower posterior margin of the rib cage and the horizontal gluteal fold), which has persisted for more than the past 3 months AND has resulted in pain on more than half the days in the past 6 months. Participants were recruited through Facebook ads across the United States and targeted emails of prior Eureka participants. As our target population was non-specific cLBP, 492 participants were excluded from the current analysis if they were pregnant, currently diagnosed with cauda equina syndrome, had severe leg weakness due to lower back pain, diagnosed with a vertebral fracture in the previous 6 months, had cancer that metastasized had or spread to bones, had cancer treatment in the last 24 months or planned in the next 12 months, or had a history of autoimmune disorders (ankylosing spondylitis, rheumatoid or psoriatic arthritis, polymyalgia rheumatica, or lupus (Figure 1).

### Measurements

The online baseline survey included questions on demographics, back-related pain, back pain treatment, medications, pain impact on quality of life, pain beliefs, medical history, health habits, and traumatic experiences. Detailed methods for all measurements used in this analysis are in Appendix A.

#### Outcome and Exposures

We selected the baseline PEG score, a three-item scale for assessing pain intensity and interference (40), as the outcome measure. We selected five exposures that have been identified in MR studies as modifiable causes of cLBP that were also measured in our dataset: alcohol use, smoking, sleep disturbance, depression, and obesity (10, 23–25, 38). Sleep disturbance and depression were measured as continuous variables using the PROMIS sleep disturbance 6a T-score (41) and PROMIS depression 4a T-score (41). The number of drinks per week, as a continuous variable, was used to measure alcohol use. Smoking (42) and obesity (43) were both analyzed as categorical variables as detailed in Appendix – Section A.

#### Confounders

Confounders of the relationships between each exposure of interest and PEG were identified using the rigorous evidence synthesis for constructing directed acyclic graphs (ESC-DAG) approach (44). This is a method for building directed acyclic graphs (DAGs) based on causal criteria which offers a structured protocol for DAG construction and reporting. DAGs are conceptual tools that are widely used to develop analytic strategies (45), especially for controlling for potential confounders in observational data analysis (8). The basic components of a DAG are nodes and edges representing variables and assumptions about their directed interrelationships. Each DAG has an exposure(s), an outcome, and a number of covariates. Differentiating confounders from other covariates, such as mediators, is critical for identifying appropriate adjustment sets to estimate statistics of interest while minimizing bias (46). The ESC-DAG method defines a systematic approach to DAG construction, by incorporating an evidence synthesis protocol into a causal inference framework, specifying how background knowledge is used for determining which variables and connections between variables are included. We followed the three step ESC-DAG method and report our decisions along with relevant literature in Appendix – Section B.

##### 1. Mapping

To begin the graph, a directed edge was drawn from each exposure to the outcome in a single graph. Next, variables collected in the BACKHOME study with a plausible association between at least one of the exposures and/or with PEG, based on the BACPAC theoretical model (1), were added as nodes. Variables that were clearly mediators of pain response (e.g. variables related to neurophysiologic pain mechanism) were excluded as we were primarily interested in estimating the total effect of our exposures. A saturated graph was created by drawing edges from each node to all the other nodes. As the resulting saturated graph was overly complex some nodes were combined if they were conceptually related and had similar inputs and outputs.

##### 2. Translation

Each relationship in the saturated graph was assessed by two authors (PZ and CO) using levels of evidence based on causal criteria (expert opinion, association, temporality, confounding control). The levels of evidence were:

a. Level 1- Expert opinion only (based on causal models published in the literature)
b. Level 2- Cross-sectional association
c. Level 3- Temporal order (longitudinal studies demonstrating that the exposure precedes cLBP)
d. Level 4- Analysis-based confounding control
e. Level 5- Design based confounding control (e.g. MR, twin studies)

All edges that included supported by Level 4 or Level 5 evidence were retained. Selected edges with lower levels of evidence that the reviewing authors (PZ and CO) felt were supported by strong theory or expert opinion were also retained. The level of evidence and supporting references for the retained edges in the saturated graph was recorded in a decision log (Appendix – Section B).

##### 3. Integration

Directed edges defined during the translation phase were synthesized into a final DAG that was used to guide the statistical analysis.

The ESC-DAG process resulted in a single, fully specified DAG that considered the five exposures simultaneously along with nodes and directed edges identified via the process of mapping, translation, and integration described above. The nodes retained following the ESC-DAG process are identified in the final DAG (Figure 1). The confounding variables retained in the DAG were age, sex, education, relationship status, financial strain, PROMIS anxiety, fear avoidance, and pain catastrophizing, as defined in the Appendix – Section A. These variables were designated Type A confounders.

BACKHOME variables that did not meet the criteria for Type A confounders, but based on evidence in the literature are plausibly associated with PEG and one or more exposures, were designated Type B confounders. Identifying potential confounders with weaker evidence allowed a sensitivity analysis for each exposure, to determine if the magnitude and direction of the effects we identified would differ substantially if these factors were adjusted for. Including these potential confounders can address one limitation of a DAG-based analysis, which is the omission of important factors needed for adjustment. However, including potential confounders with weaker evidence introduces another potential bias, as the estimate of the total effect of an exposure may be attenuated by mistakenly conditioning on a collider or mediator. The Type B confounding variables included in the sensitivity analysis were current opioid use, expectation of pain relief, post-traumatic stress disorder (PTSD), seeking compensation (lawsuit, worker’s compensation or disability claim), racial/ethnic discrimination, history of low back surgery, pain duration, self-efficacy, cognitive function, fatigue, and social isolation. These variables, with the exposures they are plausibly with, are defined in Appendix – Section A. The sensitivity analysis is reported in Appendix – Section C and described further in the analysis methods below.

### Analysis Methods

Baseline characteristics for subjects were reported as means and standard deviations (SDs) for continuous variables and counts and percentages for categorical variables. The association between our primary outcome PEG and each of the five exposure variables was estimated separately using a set of multiple linear regression models (MLRs) via the regression coefficient for the exposure of interest. Based on the comprehensive DAG identified in Figure 1, which includes all exposures along with a set of nodes and directed edges, a minimally sufficient adjustment set (MSAS) was identified for the total effect of each exposure variable separately using the R package daggity (v. 3.1) (47). The MSAS for each exposure set were included as adjustment variables in the MLR to reduce confounding bias in the estimated associations between the PEG outcome and each exposure. The result of this process is a unique adjustment set for each exposure. Two regression coefficients were estimated for each exposure via the MLRs: (1) unadjusted estimates of the exposure regression coefficient which do not control for any confounders, and (2) adjusted estimates of the exposure regression coefficient which control for the MSAS for each exposure. Given our focused examination of exposures with strong evidence in the MR literature and the objective of triangulating evidence, we focus on presenting effect estimates and confidence intervals rather than formal hypothesis testing and thus we refrain from enacting any multiplicity corrections to account for the inspection of multiple exposures. A sensitivity analysis was performed for each exposure-specific model by supplementing the exposure-specific MSAS with an additional set of adjustment factors. Results from the sensitivity analysis are reported in Appendix - Section C. All analyses were performed with SAS software (version 9.4, SAS Institute Inc., Cary, NC, USA).

## Results

Table 1 reports the baseline characteristics in the analysis study cohort.

**Table 1.** Baseline Characteristics of Vanguard Cohort.

| Baseline Characteristic | All participants<br>(n = 1376) |
| --- | --- |
| **Outcome** |  |
| PEG score (range:0-10, higher=more pain interference) | 4.4 ± 2.2 |
| **Exposures** |  |
| PROMIS sleep disturbance 6a T-score (range:31.7-76.1, higher=more disturbance) | 54.8 ± 8.0 |
| PROMIS depression 4a T-score (range:41.0-79.4, higher=more depressed) | 52.6 ± 10.1 |
| BMI |  |
| Underweight (<18.5 kg/m<sup>2</sup>) | 12 (0.9) |
| Normal weight (18.5-24.9 kg/m<sup>2</sup>) | 409 (29.8) |
| Overweight (25-29.9 kg/m<sup>2</sup>) | 410 (29.9) |
| Obesity class I (30-34.9 kg/m<sup>2</sup>) | 279 (20.3) |
| Obesity class II (35-39.9 kg/m<sup>2</sup>) | 137 (10.0) |
| Obesity class III (≥40 kg/m<sup>2</sup>) | 126 (9.2) |
| Number of alcoholic drinks per week | 2.5 ± 4.6 |
| Smoking status |  |
| Current smoker | 93 (6.8) |
| Past smoker | 457 (33.2) |
| Never smoked | 826 (60.0) |
| **Type A Confounders** |  |
| Age (years) | 54.9 ± 14.4 |
| Female | 927 (67.4) |
| Education |  |
| Some high school | 20 (1.5) |
| High school completed | 237 (17.2) |
| Associates/Technical degree completed | 208 (15.1) |
| College/Baccalaureate degree completed | 434 (31.5) |
| Doctoral/Postgraduate education | 477 (34.7) |
| Relationship status |  |
| Married | 788 (57.2) |
| Never married | 210 (15.3) |
| Divorced | 181 (13.2) |
| Domestic partner (living with partner as if married) | 100 (7.3) |
| Widowed | 77 (5.6) |
| Separated | 20 (1.5) |
| High financial strain | 145 (10.6) |
| PROMIS anxiety 4a T-score (range:40.3-81.6, higher=more anxiety) | 54.1 ± 10.1 |
| Fear avoidance score (range:0-24, higher=more avoidance) | 11.6 ± 5.9 |
| Pain catastrophizing score (range:0-12, higher=more catastrophizing) | 4.5 ± 2.6 |
| **Type B Confounders** |  |
| Current opioid use | 156 (11.3) |
| Expectation of pain relief over next 3 months (1=no relief, 10=complete relief) | 4.2 ± 2.2 |
| PTSD diagnosis, childhood and/or adult | 235 (17.1) |
| Filed workers compensation, lawsuit, or disability due to back problem or pain | 217 (15.8) |
| How often treated unfairly due to ethnicity or race |  |
| Never | 1002 (72.8) |
| Sometimes | 349 (25.4) |
| Often | 17 (1.2) |
| Always | 8 (0.6) |
| History of low-back operation |  |
| None | 1189 (86.4) |
| Decompression | 114 (8.3) |
| Spinal fusion | 73 (5.3) |
| Duration of low-back pain as ongoing problem |  |
| 3 – 6 months | 46 (3.4) |
| 6 months – 1 year | 107 (7.8) |
| 1 – 5 years | 463 (33.7) |
| More than 5 years | 759 (55.2) |
| Pain self-efficacy score (range:0-24, higher=more confidence) | 16.2 ± 5.5 |
| PROMIS cognitive function 2a T-score (range:29.5-61.2, higher=more cognitive function) | 49.2 ± 7.4 |
| PROMIS fatigue 4a T-score (range:33.7-75.8, higher=more fatigued) | 57.1 ± 10.5 |
| PROMIS social isolation 4a T-score (range:34.8-74.2, higher=more isolated) | 51.6 ± 8.2 |

Data presented as mean ± SD or n (%)

Table 2 reports the unadjusted and adjusted mean difference in baseline PEG for a given change in baseline exposure levels. Exposure effects are reported as the expected difference in baseline PEG with reference to the baseline reference category for categorical exposures or the expected difference in baseline PEG for each SD shift in the exposure for continuous exposures. The 95% confidence intervals for all of mean differences excluded zero except for the underweight BMI category, in both the unadjusted and adjusted models, and smoking status, in the adjusted model. Each SD increase in the PROMIS sleep disturbance T-score resulted in a mean 0.77 (95% CI: 0.66, 0.88) point increase in baseline PEG score in the adjusted model. In the adjusted model, each SD increase in the PROMIS depression T-score was associated with a mean 0.28 (95% CI: 0.17, 0.40) point increase in baseline PEG score. Compared to participants with normal BMI, adjusted mean PEG score was slightly higher by 0.37 points (95% CI: 0.09, 0.65) for overweight participants, about 0.8 to 0.9 points higher for those in obesity classes I and II, and 1.39 (95% CI: 0.98, 1.80) points higher for the most obese participants. Each SD decrease in number of alcoholic drinks per week resulted in a mean 0.12 (95%CI: 0.01, 0.23) increase in baseline PEG score in the adjusted model. Full results for the sensitivity analysis are presented in Appendix C. The sensitivity analysis adjusted for a wider range of factors, and the results were generally attenuated though the directions of association remained the same, and sleep disturbance and obesity remained the exposures with the strongest associations with PEG.

**Table 2.** Mean difference in baseline PEG for given change in baseline exposure.

| Exposure | Unit/referent* | Unadjusted mean difference (95% CI) | Adjusted mean difference (95% CI) |
| --- | --- | --- | --- |
| Smoking status | Never smoked |  |  |
| Current smoker |  | 1.24<br>(0.78, 1.70) | 0.36<br>(-0.07, 0.78)<sup>a</sup> |
| Past smoker |  | 0.10<br>(-0.15, 0.34) | -0.09<br>(-0.31, 0.13)<sup>a</sup> |
| PROMIS sleep disturbance T-score | +8.0 | 0.87<br>(0.76, 0.97) | 0.77<br>(0.66, 0.88)<sup>b</sup> |
| PROMIS depression T-score | +10.1 | 0.83<br>(0.72, 0.94) | 0.28<br>(0.17, 0.40)<sup>c</sup> |
| BMI | Normal weight<br>(18.5-24.9 kg/m<sup>2</sup>) |  |  |
| Underweight (<18.5 kg/m<sup>2</sup>) |  | 0.29<br>(-0.91, 1.50) | 0.29<br>(-0.86, 1.45)<sup>d</sup> |
| Overweight (25-29.9 kg/m<sup>2</sup>) |  | 0.36<br>(0.08, 0.65) | 0.37<br>(0.09, 0.65)<sup>d</sup> |
| Obesity class I (30-34.9 kg/m<sup>2</sup>) |  | 1.01<br>(0.69, 1.33) | 0.86<br>(0.55, 1.17)<sup>d</sup> |
| Obesity class II (35-39.9 kg/m<sup>2</sup>) |  | 1.00<br>(0.60, 1.41) | 0.76<br>(0.37, 1.15)<sup>d</sup> |
| Obesity class III (≥40 kg/m<sup>2</sup>) |  | 1.86<br>(1.45, 2.28) | 1.39<br>(0.98, 1.80)<sup>d</sup> |
| Number of alcoholic drinks per week | -4.6 | 0.29<br>(0.17, 0.40) | 0.12<br>(0.01, 0.23)<sup>e</sup> |

\*Continuous variable units approximate 1 SD increase or 1 SD decrease; for categorical variables, referent group is listed.
<sup>a</sup> adjusted for age, sex, depression, anxiety, sleep disturbance, education
<sup>b</sup> adjusted for age, sex, financial strain
<sup>c</sup> adjusted for age, sex, BMI, sleep disturbance, education, financial strain, fear avoidance, pain catastrophizing
<sup>d</sup> adjusted for age, sex, education, financial strain
<sup>e</sup> adjusted for age, sex, depression, anxiety, education

## Discussion

Our results demonstrate that several modifiable causal risk factors for cLBP identified by MR-alcohol use, sleep disturbance, depression, and obesity- are associated with PEG, after adjusting for confounding variables identified via a DAG constructed using a rigorous protocol. Contrary to MR studies, we did not find an association between smoking and PEG. For alcohol the direction of association was opposite what has been demonstrated in MR studies, as a decrease in alcohol use was associated with an increase, albeit very small, in PEG. For sleep disturbance, depression, and obesity the convergence of our findings with the results from MR studies, which have different designs and biases, strengthen the evidence for causal relationships between these risk factors and cLBP (7). In addition, by analyzing a cLBP cohort and using a continuous variable, PEG, as the outcome we calculated adjusted estimates for the effect of these risk factors on subjects with cLBP. The adjusted effect estimates, presented as the expected change in the PEG outcome for every standard deviation increase or decrease in the exposure (or category shift for categorical exposures) were the largest for sleep disturbance and obesity.

The major strength of our study is the rich BACKHOME dataset, which includes information on multiple risk factors and confounders for a large number of participants. The major weakness is that the validity of the results depends on several assumptions, all of which are common to analysis-based approaches to causal inference (7): no unmeasured confounders, no measurement error in the assessed confounders, and a correctly specified DAG.

While the BACKHOME dataset contains measures of a large number of potential confounders a fundamental limitation of relying on statistical adjustment for confounding variables is that unmeasured confounders can never be excluded (8). There are several potential confounding variables that are not measured in the BACKOME dataset. Some are evident from MR studies; notably, diet (16), systemic inflammation (19), physical activity (32, 36); the microbiome (11), lipids (12), personality traits (13), and blood pressure (27). Structural spinal pathology is another potential unmeasured confounder. Measurement error in the assessed confounders is a much lesser concern, given that all instruments are validated tools widely used in cLBP research.

DAGs depict the assumptions about underlying relationships between variables, which must be true in order for the research conclusions to be valid (48). RCT’s can provide strong evidence for causal relationships, while the evidence from other study designs is necessarily weaker. The Austin Bradford Hill considerations (49), a framework based on inductive reasoning, is commonly used to assess causality, but the only universally agreed upon criterion from that framework is temporality (i.e. cause precedes effect) (49). In the absence of RCT’s there is no consensus on the grading of evidence for causal relationships. As a result, the assumptions underlying DAGs generally rely heavily on judgements by domain experts (8). In fact, a recent review found that only 6% of published DAG’s provided citations supporting one or more edges between nodes (8). A particular strength of our study is that the ESC-DAG method we followed for DAG construction combines methodological rigor-including elements from the Hill considerations and contemporary causal inference methods as well as expert opinion-with detailed documentation. Nevertheless, as with all DAGs, there are built-in assumptions which cannot be proven.

A major limitation of our study is that the data are cross sectional. Therefore, while prior evidence, as documented in decision log in Appendix B, supports the temporal relationships depicted in our DAG, reverse causation cannot be excluded. This is a particular concern for risk factors where bidirectional causal relationships have been demonstrated (13). Although commonly referred to as “feedback loops”, these relationships actually represent co-evolution of variables over time, with the current state of one variable impacting the future state of another variable, which may in turn affect the future state of the original variable (50). A simplified depiction of a bi-directional relationship with depression and PEG is in figure 3.

**Figure 2:**
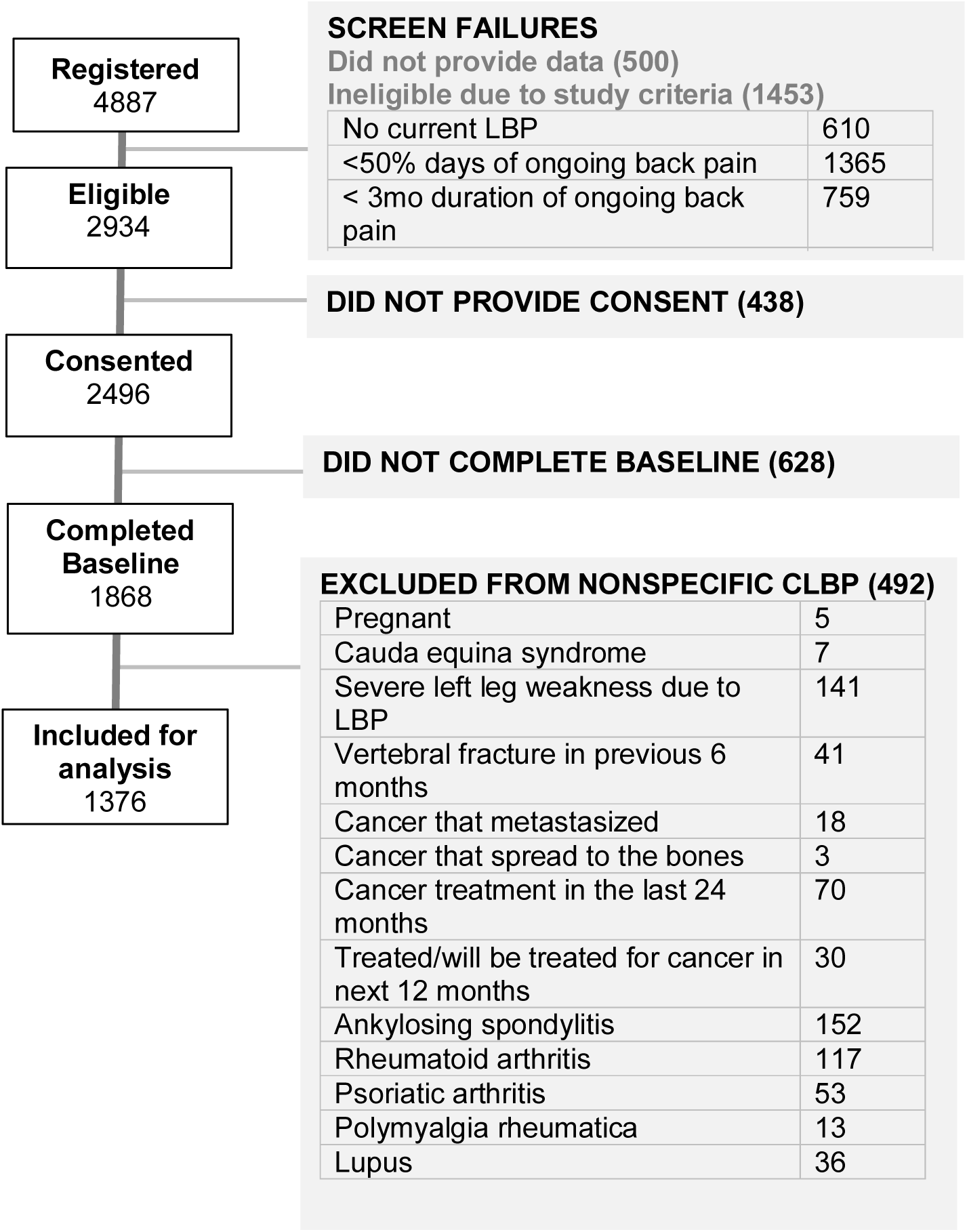

**Figure 3.**
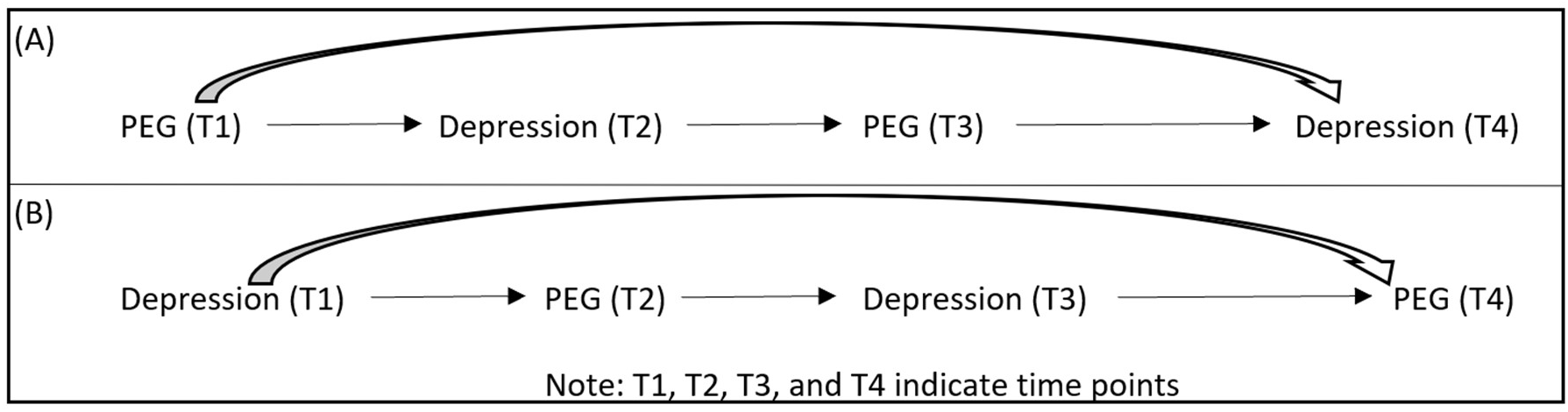
Directed acyclic (DAG) representations of the bidirectional effects over time. (A)PEG is exposure of interest and depression is outcome of interest; (B) Depression is exposure of interest and PEG is outcome interest. Adapted from Kunicki EJM, Zach. OSF preprints | As the Wheel Turns: Causal inference for Feedback Loops and Bidirectional Effects. 2024 (49).

Time varying relationships can also extend to both confounding variables as well as mediators (variables that are in the causal pathway from the exposure to the outcome). As longitudinal data becomes available we will be able to assess the effects of time-varying relationships on our results.

Despite the limitations inherent in our study design, the triangulation of evidence from our analysis-based approach with the results from design-based MR approaches supports causal links between cLBP and three of the risk factors we studied-sleep disturbance, depression, and obesity. The key assumptions about MR studies are that the genetic variant (which serves as an instrumental variable) is robustly associated with the exposure, is not associated with confounders, and is not associated with the outcome other than via its association with the exposure (7). The biases in MR studies, then, are different than the sources of bias in our study. The concordance between MR studies and our findings strengthens the evidence that sleep disturbance, depression, and obesity are causal risk factors for cLBP, because the chance that studies with very different potential sources of bias would align to give similar results is presumably small (7). The evidence from MR studies on the association between alcohol use and cLBP is mixed, with one study showing an association (51), and another not (24). The study by Lv, et al. (24) measured alcohol consumption as the number of drinks per week, as we did, while the study by Williams, et al (38) measured the frequency of alcohol intake, defined as a categorical variable. The different measurements for alcohol consumption may account for the different findings in MR studies. The evidence from MR studies on the association between smoking and cLBP is consistent, although with relative small odds ratios (OR’s), varying between 1.36 (24) and 1.27 (38). While the evidence from MR studies supports a causal association between smoking and cLBP the lack of convergence with our findings suggests further study is needed.

In MR studies causal effect estimates are reported as odds ratios (OR’s), using a case definition of cLBP as the outcome. Our results complement these findings by estimating of the association of change in PEG with reductions in exposures. Estimating the effects of exposures on the absolute scale of PEG, as opposed to a relative measure of association like the OR, is more meaningful for choosing interventions. The effect sizes for the exposures we studied are generally small, below the minimally important difference (MID) for PEG of 1.0 (52). However, these are average effects for the population, and in individual patients the effects of an exposure may be greater. Furthermore, in any one individual there may be a collection of component causes, each of which must be present for an outcome to occur, a concept known as the sufficient cause framework (49). Previously, interventions for the risk factors we studied have focused on each one individually (51, 53–56), with generally disappointing results. A more effective approach may be individualized, multimodal treatment plans that address all causal risk factors. Addressing sleep disturbance and obesity, which have the greatest effect sizes in our study, may be particularly important. There are a number of evidence-based treatments for insomnia, which could be incorporated into a multimodal cLBP treatment program (55). Interventions for obesity in cLBP patients have focused on lifestyle interventions (51), but weight loss drugs for those patients that fit the indications may be a more effective strategy.

In summary, in this study we analyzed baseline data from a unique cLBP cohort, which includes information on multiple risk factors and confounders for a large number of subjects. Using rigorous confounding control we found associations between alcohol use, sleep disturbance, depression, and obesity and PEG. Convergence of our findings for sleep disturbance, depression, and obesity with the results from MR studies, which have different designs and biases, strengthens the evidence that these factors are not just associated with cLBP but cause cLBP. As longitudinal data becomes available from the cohort we will be able to assess the effects of time-varying relationships on our results. The effect of reducing each of these risk factors on PEG was small, with the greatest effects associated with sleep disturbance and obesity. The effect of incorporating treatment of the risk factors we have identified into multimodal cLBP treatment strategies should be a focus of future study.

## Data Availability

All data produced in the present study are available upon reasonable request to the authors

## Appendix A: Study measurement details

### Outcome measure

The **PEG score** ranges from 0-10, with higher values indicating more pain interference. It is calculated as the mean of responses to 3 items:

1. Average pain in past week (0=no pain, 10=worst imaginable pain)
2. How much pain interfered with enjoyment of life in past week (0=did not interfere, 10=completely interfered)
3. How much pain interfered with general activity in past week (0=did not interfere, 10=completely interfered)

### Exposures

The **PROMIS sleep disturbance** 6a t-score (range 31.7-76.1, higher=more sleep disturbance) was based on responses to the following 6 questions:

1. In the past 7 days, my sleep quality was: 1 (=very good) to 5 (=very poor)
2. In the past 7 days, my sleep was refreshing: 1 (=very much) to 5 (=not at all) Responses to 3-6 ranged from 1 (=not at all) to 5 (=very much) In the past 7 days:
3. I had a problem with my sleep.
4. I had difficulty falling asleep.
5. My sleep was restless.
6. I tried hard to get to sleep.

The responses to these questions were summed to produce a raw summary score (range 6-30), which was then mapped to a t-score, with 50 representing the mean of a reference population and 10 being the SD of that population.

The **PROMIS depression** 4a t-score (range 41.0-79.4, higher=more depressed) was based on responses to the following 4 questions, with responses ranging from 1 (=never) to 5 (=always): In the past 7 days…

1. I felt worthless.
2. I felt helpless.
3. I felt depressed.
4. I felt hopeless.

The responses to these questions were summed to produce a raw summary score (range 4-20), which was then mapped to a t-score, with 50 representing the mean of a reference population and 10 being the SD of that population.

**Alcohol use** was defined as number of drinks per week as a continuous variable in response to the question: How many alcoholic drinks do you consume per week, on average?

**Smoking** was measured as a categorical variable in reponse to the question: How would you describe your cigarette smoking?

1. Never smoked
2. Current smoker
3. Used to smoke, but have now quit

**Body mass index (BMI)** was calculated from self-reported weight and height. Obesity was defined by BMI of 30.0 kg/m^2^ or higher. Furthermore, underweight was defined as <18.5 kg/m^2^, normal weight ranges from 18.5-24.9 kg/m^2^, overweight ranges from 25-29.9 kg/m^2^, obesity class I ranges from 30-34.9 kg/m^2^, obesity class II ranges from 35-39.9 kg/m^2^, and obesity class III constitutes ≥40 kg/m^2^.

### Type A confounding variables

**Age** was measured as a continuous variable and **sex** as a binary variable (male/female). Categories for **education** were some high school, high school completed, associates/technical degree completed, college/baccalaureate degree completed, doctoral/postgraduate education. Categories for **relationship status** were married, never married, divorced, domestic partner, widowed, separated. Participants were asked how difficult it was to pay for basic necessities; “hard” and “very hard” responses were classified as high **financial strain**.

The **PROMIS anxiety 4a T-score** (range:40.3-81.6, higher=more anxiety) was based on responses to the following 4 questions, with responses ranging from 1 (=never) to 5 (=always): In the past 7 days…

1. I felt fearful
2. I found it hard to focus on anything other than my anxiety
3. My worries overwhelmed me
4. I felt uneasy

The responses to these questions were summed to produce a raw summary score (range 4-20), which was then mapped to a t-score, with 50 representing the mean of a reference population and 10 being the SD of that population.

The **Fear Avoidance score** (FABQ-PA) (57) (range 0-24, higher=more avoidance) was calculated as the sum of responses (0=completely disagree, 6=completely agree) to the following 4 items:

1. Physical activity makes my pain worse.
2. Physical activity might harm my back.
3. I should not do physical activities which might make my pain worse.
4. I cannot do physical activities which might make my pain worse.

The **Pain Catastrophizing Scale** SF (PCS-6) (58) (range 0-12, higher=more catastrophizing) included 3 subscales -- Helplessness, Magnification, and Rumination -- with each subscale having 2 components. The responses to the following 6 statements ranged from 0=not at all to 4=all the time:

When I’m in pain…

1. It’s awful and I feel that it overwhelms me. (Helplessness subscale)
2. I feel I can’t stand it anymore. (Helplessness subscale)
3. I become afraid that the pain will get worse. (Magnification subscale)
4. I wonder whether something serious may happen. (Magnification subscale)
5. I keep thinking about how much it hurts. (Rumination subscale)
6. I keep thinking about how badly I want the pain to stop. (Rumination subscale)

The mean for each subscale was determined, resulting in a 0-4 score. The 3 subscales were then summed to create the total score, ranging from 0-12, with higher scores representing more pain catastrophizing.

### Type B confounding variables

**Current opioid use** was defined as current use for low-back pain or current daily use for any reason. **Expectation of pain relief** over next 3 months was assessed by the question: Please indicate how much pain relief you expect over the coming three months (range 1 = no relief, and 10 = complete relief). **Duration of low back pain** was recorded as 3-6 months, 6 months −1 year, 1-5 years or more than 5 years. **History of low-back operation** was assessed and recorded as none, decompression surgery or spinal fusion surgery. **Discrimination** was assessed by the question: How often do people treat you unfairly because of your ethnicity or race? 1= never, 2 = sometimes, 3 = often, 4 = always.

**Post traumatic stress disorder (PTSD)** was noted if participant marked “yes” to having experienced things as a child or as an adult that are unusually or especially frightening, horrible, or traumatic (examples include a serious accident or fire, a physical or sexual assault or abuse, an earthquake or flood, a war, seeing someone be killed or seriously injured, having a loved one die through homicide or suicide) AND participant reported at least 3 of 5 symptoms in past month (nightmares, avoided triggering situations, on guard, felt detached, self-blamed).

Those who marked “yes” to “have you filed or been awarded a worker’s compensation claim related to your back problem,” “are you involved in a lawsuit or legal claim related to your back problem,” or “have you ever applied for, or received, disability insurance for your pain condition” were marked as having filed **workers compensation, lawsuit, or disability due to back problem or pain**.

**Pain self-efficacy score** (PSEQ-4) (59) was calculated as the sum of responses to 4 questions on how confident the participant was in doing the following (0=not at all confident, 6=completely confident):

1. I can cope with my pain in most situations.
2. I can still do many of the things I enjoy doing, such as hobbies or leisure activity, despite the pain.
3. I can still accomplish most of my goals in life, despite the pain.
4. I can live a normal lifestyle, despite the pain.

The total score ranged from 0-24, with higher scores representing greater confidence.

T-scores were calculated for the following PROMIS measures: **cognitive function** 2a, **fatigue** 4a, and **social isolation** 4a (41).

## Appendix B: ESC-DAG decision log

### Decision Log Type A confounders

| **Edge** | **Level of Evidence<sup>a</sup>** | **References** |
| --- | --- | --- |
| Anxiety -> Comorbidity (CCI) | 5 | (60-63) |
| Depression -> Comorbidity (CCI) | 5 | (60-68) |
| Sleep -> Comorbidity (CCI) | 5 | (69-72) |
| Financial strain -> Obesity | 3 | (60, 73-75) |
| Obesity -> Comorbidity (CCI) | 5 | (60, 76) |
| Obesity -> Depression | 5 | (60) |
| Obesity -> Anxiety | 5 | (60, 76, 77) |
| Obesity -> PEG | 5 | (10, 33, 60) |
| Anxiety -> Sleep | 5 | (72, 78, 79) |
| Anxiety -> Smoking | 5 | (78, 80, 81) |
| Depression -> Smoking | 5 | (78, 80, 81) |
| Sleep -> Depression | 5 | (72, 78, 82) |
| Depression -> PEG | 5 | (31, 38, 60, 83, 84) |
| Anxiety -> PEG | 5 | (10, 60, 84) |
| Sleep -> PEG | 5 | (25, 31, 78) |
| Education -> Comorbidity (CCI) | 5 | (85-89) |
| Education -> Obesity | 5 | (89, 90) |
| Education -> Anxiety | 5 | (63, 91) |
| Education -> Depression | 5 | (63, 78, 89, 91, 92) |
| Education -> Financial stress | 3 | (93, 94) |
| Education -> Smoking | 5 | (83, 95, 96) |
| Education -> PEG | 5 | (38, 78, 97, 98) |
| Financial strain -> Anxiety | 3 | (99, 100) |
| Financial strain -> Depression | 3 | (99-101) |
| Financial strain -> Sleep | 2 | (102, 103) |
| Financial strain -> PEG | 2 | (78, 97, 104) |
| Sleep -> Smoking | 5 | (72, 78) |
| Smoking -> PEG | 5 | (24, 38, 98) |
| Catastrophization -> Anxiety | 2 | (105, 106) |
| Catastrophization -> Depression | 2 | (105, 106) |
| Catastrophization -> PEG | 4 | (84, 105, 107-111) |
| Fear Avoidance -> Anxiety | 2 | (105, 106, 112) |
| Fear Avoidance -> Depression | 2 | (105, 106, 112) |
| Fear Avoidance -> PEG | 4 | (84, 105, 109-111) |
| Age -> Comorbidity (CCI) | 2 | (60, 113) |
| Age -> Obesity | 3 | (60, 114) |
| Age -> Sleep | 3 | (78, 115) |
| Age -> PEG | 4 | (60, 78, 105, 116, 117) |
| Age -> Smoking | 3 | (78, 118) |
| Age -> Anxiety | 3 | (78, 105, 119-122) |
| Age -> Depression | 3 | (78, 105, 119, 123-125) |
| Age -> Catastrophization | 2 | (105, 112, 126) |
| Age -> Fear Avoidance | 2 | (105, 112, 127) |
| Age -> Financial Strain | 2 | (128) |
| Sex -> Comorbidity (CCI) | 5 | (60, 129) |
| Sex -> Obesity | 5 | (60, 129) |
| Sex -> Sleep | 3 | (130) |
| Sex -> PEG | 4 | (60, 98, 131) |
| Sex -> Smoking | 3 | (132) |
| Sex -> Anxiety | 3 | (60, 133) |
| Sex -> Depression | 3 | (60, 133) |
| Sex -> Catastrophization | 3 | (105, 126, 131) |
| Sex -> Fear Avoidance | 3 | (105, 134) |
| Sex -> Financial strain | 4 | (135) |
| Sex -> Education | 3 | (105, 136, 137) |
| Alcohol -> Sleep | 5 | (138) |
| Anxiety -> Alcohol | 5 | (139-141) |
| Depression -> Alcohol | 5 | (142-145) |
| Age -> Alcohol | 5 | (146-148) |
| Sex -> Alcohol | 5 | (148) |
| Alcohol -> Financial strain | 5 | (144) |
| Education -> Alcohol | 3 | (149) |
| Alcohol -> Comorbidity (CCI) | 3 | (148, 150) |
| Alcohol -> Obesity | 5 | (151) |

<sup>a</sup>Levels of evidence:

Level 1- Expert opinion only (based on causal models published in the literature)

Level 2- Cross-sectional assocation

Level 3- Temporal order (longitudinal studies demonstrating that the exposure precedes cLBP)

Level 4- Analysis-based confounding control

Level 5- Design based confounding control (e.g. MR, twin studies)

### Decision Log Type B confounders

| **Confounder** | **Exposures** | **References** |
| --- | --- | --- |
| Expectations | Obesity | (152-157) |
| Self Efficacy | Obesity, Alcohol, Smoking | (84, 110, 158-164) |
| PTSD | Depression, Obesity, Sleep, Smoking, Alcohol | (165-171) |
| Compensation | Obesity, Smoking, Alcohol | (172-182) |
| Mistreatment | Depression, Obesity, Sleep, Smoking, Alcohol | (183-190) |
| Relationship status | Depression, Obesity, Sleep, Smoking, Alcohol | (191-196) |
| Surgery | Depression, Obesity, Sleep, Smoking, Alcohol | (197-204) |
| Opioid use | Depression, Obesity, Sleep, Smoking, Alcohol | (205-210) |
| Duration of LBP | Depression, Obesity, Sleep, Smoking, Alcohol | (57, 172, 211-219) |
| Isolation | Depression, Sleep, Alcohol | (220-224) |
| Fatigue | Depression, Obesity, Sleep, Smoking, Alcohol | (225-233) |
| Cognitive Function | Depression, Obesity, Sleep, Smoking, Alcohol | (234-239) |

## Appendix C: Exposure-specific sensitivity analysis

Appendix C Table 1 reports the unadjusted, adjusted, and sensitivity analysis adjusted (based on the combined ESC-DAG adjustment set supplemented with confounders from the sensitivity analysis) mean difference in baseline PEG for a given change in baseline exposure levels. Effect estimates are reported as described in the main paper for each model. Compared to participants who never smoked, the sensitivity analysis adjusted mean baseline PEG score for current smokers was about a quarter of a point higher (mean difference = 0.28; 95% CI: −0.09, 0.64). There was not a significant difference in mean PEG score for past smokers vs. never smokers. Each SD increase in the PROMIS sleep disturbance T-score resulted in a mean 0.36 (95% CI: 0.24, 0.48) point increase in baseline PEG score in the sensitivity analysis adjusted model. In the sensitivity adjusted model, there was no longer an association between depression and PEG: each SD increase in the PROMIS depression T-score was associated with a mean 0.08 (95% CI: −0.05, 0.23) point increase in baseline PEG score. Compared to participants with normal BMI, sensitivity analysis adjusted mean PEG score was slightly higher by 0.24 points (95% CI: 0.02, 0.47) for overweight participants, about 0.3 to 0.4 points higher for those in obesity classes I and II, and 0.55 (95% CI: 0.21, .89) points higher for the most obese participants. There was no association between alcohol use and PEG in the sensitivity analysis. The sensitivity analysis adjusted for a wider range of factors and the results were generally attenuated though the directions of association remained the same, and sleep disturbance and obesity remained the exposures with the strongest associations with PEG.

**Appendix C Table 1.**
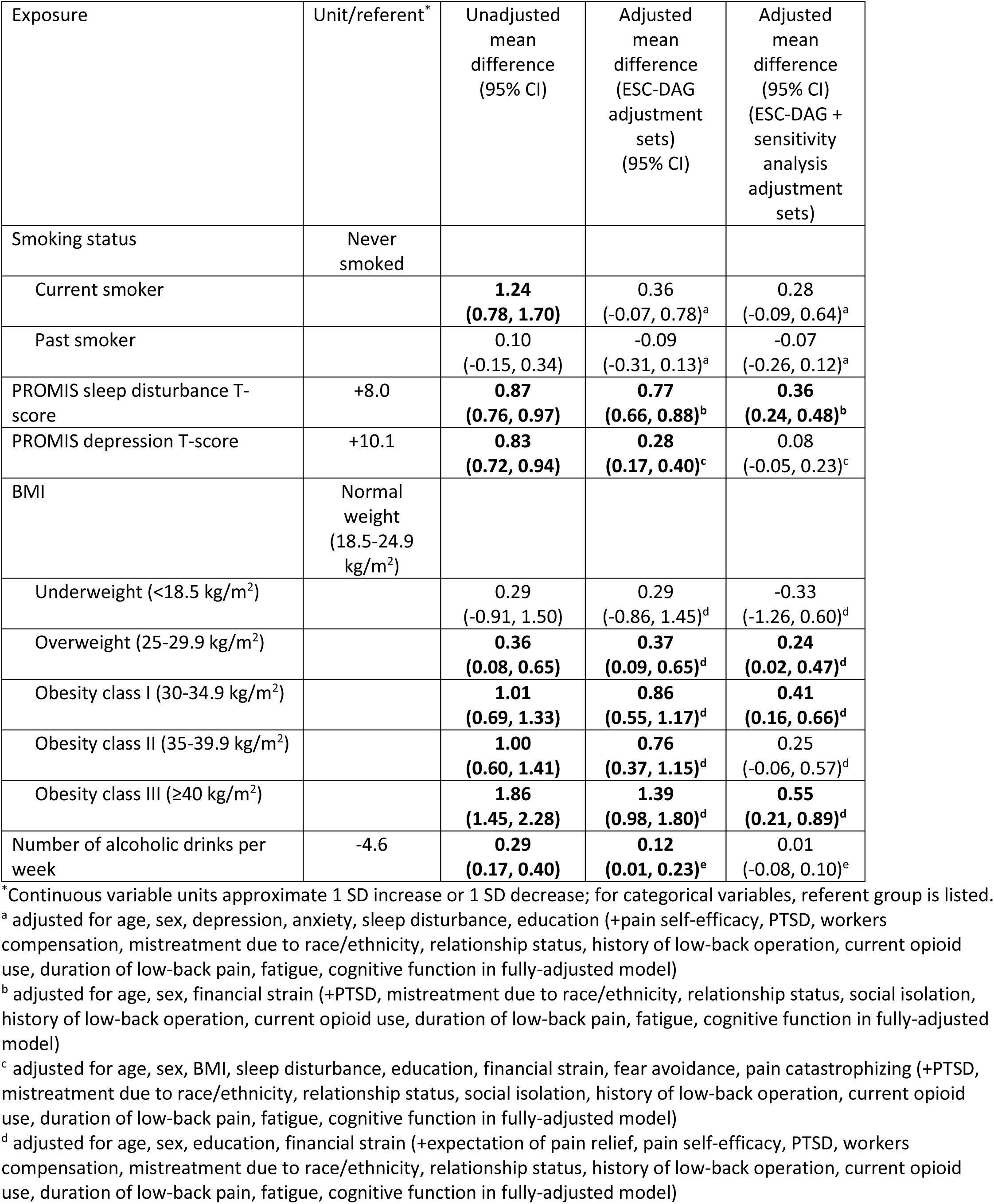
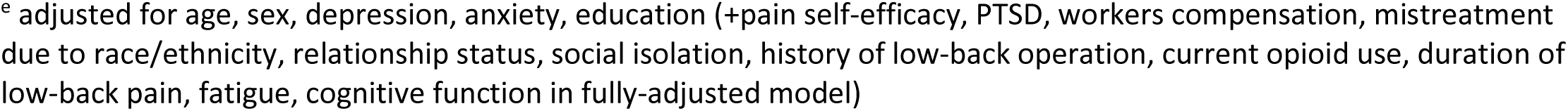
Mean difference in baseline PEG for given change in baseline exposure.

| Exposure | Unit/referent* | Unadjusted mean difference (95% CI) | Adjusted mean difference (ESC-DAG adjustment sets) (95% CI) | Adjusted mean difference (95% CI) (ESC-DAG + sensitivity analysis adjustment sets) |
| --- | --- | --- | --- | --- |
| Smoking status | Never smoked |  |  |  |
| Current smoker |  | **1.24**<br>**(0.78, 1.70)** | 0.36<br>(-0.07, 0.78)<sup>a</sup> | 0.28<br>(-0.09, 0.64)<sup>a</sup> |
| Past smoker |  | 0.10<br>(-0.15, 0.34) | -0.09<br>(-0.31, 0.13)<sup>a</sup> | -0.07<br>(-0.26, 0.12)<sup>a</sup> |
| PROMIS sleep disturbance T-score | +8.0 | **0.87**<br>**(0.76, 0.97)** | **0.77**<br>**(0.66, 0.88)<sup>b</sup>** | **0.36**<br>**(0.24, 0.48)<sup>b</sup>** |
| PROMIS depression T-score | +10.1 | **0.83**<br>**(0.72, 0.94)** | **0.28**<br>**(0.17, 0.40)<sup>c</sup>** | 0.08<br>(-0.05, 0.23)<sup>c</sup> |
| BMI | Normal weight (18.5-24.9 kg/m<sup>2</sup>) |  |  |  |
| Underweight (<18.5 kg/m<sup>2</sup>) |  | 0.29<br>(-0.91, 1.50) | 0.29<br>(-0.86, 1.45)<sup>d</sup> | -0.33<br>(-1.26, 0.60)<sup>d</sup> |
| Overweight (25-29.9 kg/m<sup>2</sup>) |  | **0.36**<br>**(0.08, 0.65)** | **0.37**<br>**(0.09, 0.65)<sup>d</sup>** | **0.24**<br>**(0.02, 0.47)<sup>d</sup>** |
| Obesity class I (30-34.9 kg/m<sup>2</sup>) |  | **1.01**<br>**(0.69, 1.33)** | **0.86**<br>**(0.55, 1.17)<sup>d</sup>** | **0.41**<br>**(0.16, 0.66)<sup>d</sup>** |
| Obesity class II (35-39.9 kg/m<sup>2</sup>) |  | **1.00**<br>**(0.60, 1.41)** | **0.76**<br>**(0.37, 1.15)<sup>d</sup>** | 0.25<br>(-0.06, 0.57)<sup>d</sup> |
| Obesity class III (≥40 kg/m<sup>2</sup>) |  | **1.86**<br>**(1.45, 2.28)** | **1.39**<br>**(0.98, 1.80)<sup>d</sup>** | **0.55**<br>**(0.21, 0.89)<sup>d</sup>** |
| Number of alcoholic drinks per week | -4.6 | **0.29**<br>**(0.17, 0.40)** | **0.12**<br>**(0.01, 0.23)<sup>e</sup>** | 0.01<br>(-0.08, 0.10)<sup>e</sup> |

\*Continuous variable units approximate 1 SD increase or 1 SD decrease; for categorical variables, referent group is listed.
<sup>a</sup> adjusted for age, sex, depression, anxiety, sleep disturbance, education (+pain self-efficacy, PTSD, workers compensation, mistreatment due to race/ethnicity, relationship status, history of low-back operation, current opioid use, duration of low-back pain, fatigue, cognitive function in fully-adjusted model)
<sup>b</sup> adjusted for age, sex, financial strain (+PTSD, mistreatment due to race/ethnicity, relationship status, social isolation, history of low-back operation, current opioid use, duration of low-back pain, fatigue, cognitive function in fully-adjusted model)
<sup>c</sup> adjusted for age, sex, BMI, sleep disturbance, education, financial strain, fear avoidance, pain catastrophizing (+PTSD, mistreatment due to race/ethnicity, relationship status, social isolation, history of low-back operation, current opioid use, duration of low-back pain, fatigue, cognitive function in fully-adjusted model)
<sup>d</sup> adjusted for age, sex, education, financial strain (+expectation of pain relief, pain self-efficacy, PTSD, workers compensation, mistreatment due to race/ethnicity, relationship status, history of low-back operation, current opioid use, duration of low-back pain, fatigue, cognitive function in fully-adjusted model)
<sup>e</sup> adjusted for age, sex, depression, anxiety, education (+pain self-efficacy, PTSD, workers compensation, mistreatment due to race/ethnicity, relationship status, social isolation, history of low-back operation, current opioid use, duration of low-back pain, fatigue, cognitive function in fully-adjusted model)

## Notes

Research reported in this publication was supported by the National Institute of Arthritis and Musculoskeletal and Skin Diseases of the National Institutes of Health under Award Number U19AR076737. The content is solely the responsibility of the authors and does not necessarily represent the official views of the National Institutes of Health. The Core Center of Patient-centric, Mechanistic Phenotyping in Chronic Low Back (REACH) investigators include the following University of California, San Francisco (unless noted otherwise) personnel in alphabetical order: Zehra Akkaya, PhD Prakruthi Amarkumar, PhD Jeannie Bailey, PhD Julia Barylak Sigurd Berven, MD Andrew Bishara, MD Dennis M. Black, PhD Noah Bonnheim, PhD Atul Butte, MD, PhD Jennifer Cummings Karina Del Rosario, MD Emilia Demarchis, MD Sibel Demir-Deviren, MD Susan K. Ewing, MS Adam Ferguson, PhD Aaron Fields, PhD Scott M. Fishman, MD (University of California, Davis) Sergio Garcia Guerra Fatemeh Gholi Zadeh Kharrat, PhD Xiaojie (Summer) Guo Misung Han, PhD Trisha Hue, PhD J. Russell Huie, PhD C. Anthony Hunt, PhD Anastasia Keller, PhD Karim Khattab Roland Krug, PhD Gregorji Kurillo, PhD Feng Lin Thomas Link, MD, PhD Jeffrey Lotz, PhD John Lynch, PhD Tong Lyu Rob Matthew, PhD Wolf Mehling, MD Esmeralda Mendoza, MPH Praveen Mummaneni, MD, MBA Caroline Navy Conor O’Neill, MD Jessica Ornowski Thomas Peterson, PhD Ananya Rupanagunta (University of California, Berkeley) Aaron Scheffler, PhD, MS Shalini Shah, MD (University of California, Irvine) Irina Strigo, PhD Naoki Takegami, MD Abel Torres-Espin, PhD (University of Waterloo) Salvatore Torrisi, PhD Sachin Umrao, PhD Rohit Vashisht, PhD Joanna Veres An (Joseph) Vu, PhD Mark Steven Wallace, MD (University of California, San Diego) Lucy Ann Wu, MPH Po-Hung Wu, PhD Patricia Zheng, MD Jiamin Zhou, MS

### Competing Interest Statement

All authors have completed the ICMJE uniform disclosure form at www.icmje.org/coi_disclosure.pdf and declare: no support from any organization for the submitted work; JL owns shares of Bioniks, LLC (a company commercializing a depth camera system similar to that being used by the REACH Physical Function and Biomechanics Core), is an Officer for Bioniks and serves on the Board of Directors; JL owns shares of Aclarion and is a consultant for Aclarion (a company which has developed and is marketing the magnetic resonance spectroscopy (MRS) diagnostic tool that is being used for the REACH comeBACK patient cohort); CO owns options of Aclarion; and JB owns options of Bioniks.

### Funding Statement

Research reported in this publication was supported by the National Institute Of Arthritis And Musculoskeletal And Skin Diseases of the National Institutes of Health under Award Number U19AR076737. The content is solely the responsibility of the authors and does not necessarily represent the official views of the National Institutes of Health.

### Author Declarations

The BACKHOME study has been approved by the WCG Institutional Review Board (IRB) under reliance agreements from the UCSF, UCD, UCI, and UCSD IRBs.

### Summary of Updates

Clarified mistake in author list

